# Statistical Analysis Plan for the REDEP study: Effectiveness of dual-mobility cups for preventing dislocation after primary total hip arthroplasty by a posterolateral approach compared to unipolar cups in elderly patients

**DOI:** 10.64898/2026.10.27.26364841

**Authors:** Okke Hoonhout, Nienke W Willigenburg, Rudolf W Poolman, Esther Scheijbeler, Amy Hofman, Nini H Jonkman

## Abstract

Periprosthetic hip dislocation is a frequent complication after primary total hip arthroplasty. Observational studies suggest that dual mobility cups may reduce dislocation risk to approximately 1%, but definitive randomized evidence supporting their routine use in primary total hip arthroplasty is absent.

The REDEP trial is the first prospective, randomized controlled trial that compares dual mobility cups with unipolar cups in patients ≥70 years who electively undergo primary total hip arthroplasty via a posterolateral approach. The primary objective of the trial is to determine the difference in the number of patients who suffer a periprosthetic dislocation of the hip within 1 year after primary total hip arthroplasty in this population. Other outcomes include the difference in the total number of hip dislocations (including recurrent dislocations within patients), differences in patient-reported outcome measures, and differences in the incidence of (serious) adverse events.

This statistical analysis plan describes the primary statistical analysis for the REDEP study. The analyses described herein were defined prior to database lock and unblinding. Secondary analyses will be described in subsequent versions of this statistical analysis plan.

## **1.** Introduction

### 1.1 Background and Rationale

Periprosthetic hip dislocation is a frequent complication after primary total hip arthroplasty (THA) (3). Dual mobility acetabular cups were developed to improve hip stability by incorporating a mobile polyethylene liner between the femoral head and the acetabular shell, thereby reducing the risk of periprosthetic hip dislocation. Observational studies suggest that dual mobility cups may reduce dislocation risk to approximately 1% (4, 5, 6).

For this reason, dual mobility cups are increasingly adopted in clinical practice to reduce instability, despite the absence of definitive randomized evidence supporting their routine use in primary THA. The REDEP trial is the first prospective, randomized controlled trial that compares dual mobility cups with unipolar cups in patients ≥70 years, who electively undergo primary THA via a posterolateral approach. It is nested within the Dutch Arthroplasty Register (LROI), enabling efficient data collection on clinical outcomes, patient-reported outcomes (PROMs), and long-term implant survival.

This SAP describes the primary statistical analysis for the REDEP study. The analyses described herein were defined prior to database lock and unblinding. Secondary analyses will be described in subsequent versions of this SAP.

### 1.2 Objectives

#### Primary objective

- Determine the difference in the number of patients who suffer a periprosthetic dislocation of the hip within 1 year after primary THA via a posterolateral approach with a dual mobility cup compared to a unipolar cup in patients ≥70 years old.

#### Secondary objectives

- Determine the difference in the total number of hip dislocations (including recurrent dislocations within patients) within 1 year after primary THA via a posterolateral approach with a dual mobility cup compared to a unipolar cup in patients ≥70 years old.
- Determine the difference in PROMs 1 year after primary THA via a posterolateral approach with a dual mobility cup compared to a unipolar cup in patients ≥70 years old.
- Compare the incidence of dislocation, revision surgery, prosthetic joint infection and mortality within 1 year after primary THA via a posterolateral approach with a dual mobility cup compared to a unipolar cup in patients ≥70 years old.

#### Objectives to be detailed in later versions of this SAP

1 year outcomes

- Compare the cost-effectiveness and cost-utility of dual mobility cups compared to unipolar cups, from a healthcare and societal perspective, in patients ≥70 years old undergoing primary THA via a posterolateral approach, at 1 year follow-up.
- Determine the win odds for patients in the REDEP study based on a hierarchical composite of multiple outcomes at 1 year follow-up.

2 year outcomes

- Determine the difference in number of patients with a hip dislocation and the total number of hip dislocations (including recurrent dislocations within patients) within 2 years after primary THA via a posterolateral approach with a dual mobility cup compared to a unipolar cup in patients ≥70 years old.
- Determine the difference in PROMs 2 years after primary THA via a posterolateral approach with a dual mobility cup compared to a unipolar cup in patients ≥70 years old.
- Compare the incidence of dislocation, revision surgery, prosthetic joint infection and mortality within 2 year after primary THA via a posterolateral approach with a dual mobility cup compared to a unipolar cup in patients ≥70 years old.

5-10 year outcomes

- Compare long-term (5- and 10-year) implant survival between a dual mobility cup and a unipolar cup in patients ≥70 years old undergoing primary THA via a posterolateral approach, based on LROI registry data

## 2. Study Methods

### 2.1 Trial Design

The REDEP study is a prospective, multi-center, international, single-blinded, two-arm parallel randomized controlled trial nested in the LROI registry comparing dual mobility cups to unipolar cups in patients ≥70 years undergoing primary THA via a posterolateral approach.

### 2.2 Randomization

After providing written informed consent, patients were randomized 1:1 to the dual mobility cup group or the unipolar cup group using the Castor electronic data capture system (7). Variable randomization blocks of 2, 4 and 6 were used, stratified by hospital.

### 2.3 Sample Size

The sample size was calculated for the primary outcome and was based on anticipated 1-year dislocation risks of 4% in the unipolar cup group and 1% in the dual mobility cup group. To show the superiority of dual mobility cups compared to unipolar cups with respect to the number of patients who suffer a hip dislocation within 1 year after primary THA, with at least 80% power at a two-sided significance level of 0.05, 976 patients (488 in each group) would need to participate. To account for loss to follow-up, 550 patients are recruited per arm (1,100 total randomized).

### 2.4 Hypothesis Testing Framework

The primary analysis is a superiority test. The null hypothesis is that there is no difference in the number of patients who suffer a hip dislocation within 1 year after primary THA. The alternative hypothesis is that dual mobility cups result in a lower number of patients who suffer a hip dislocation within 1 year after primary THA. All statistical tests are two-sided with a significance level of α = 0.05, unless stated otherwise.

### 2.5 Interim Analysis

**Timing:** As per protocol, an interim analysis was performed when 200 patients reached the 3-month post-operative PROM evaluation point (see section 3.7). The interim analysis was performed in April 2021 and included data from 215 patients (101 in the dual mobility cup group and 114 in the unipolar cup group).

**Method:** In the interim analysis, the number of patients who suffered a hip dislocation was compared between the unipolar cup group and the dual mobility cup group. We used Fisher’s exact test, since the assumptions for the chi-square test were not met (one cell contained zero observed counts). The number of revisions and Serious Adverse Events (SAEs) were tabulated but not formally tested.

**Alpha adjustment:** To control the overall type I error rate associated with repeated analyses of accumulating data, the O’Brien-Fleming alpha-spending approach was applied (8). With one planned interim analysis and one final primary analysis, the two-sided significance levels are 0.0054 and 0.0492, respectively. These significance levels preserve an overall two-sided type I error rate of 0.05 (9).

**Decision:** At the interim analysis, 0 dislocations occurred in 101 patients (0%) in the dual mobility cup group, compared with 3 of 114 patients (2.6%) in the unipolar cup group (Fisher’s exact test, p = 0.249). This did not meet the pre-specified O’Brien-Fleming stopping boundary of p < 0.0054. Two patients in the unipolar cup group suffered recurrent dislocations (3 each), resulting in a total of 7 dislocations in this group. The results were discussed with the steering committee, Van Rens Foundation and the Medical Research Ethics Committee and it was decided to continue the trial.

### 2.6 Timing of Final Analysis

Q1 2026: final patients completed their 1-year follow-up.

Q2 2026: final data extracted from all relevant data sources and participating hospitals Q2-Q3 2026: verification and (pre-)processing of raw extracted data

Q3 2026: data analyses of primary and secondary outcomes included in the main manuscript

Q4 2026 and later: additional analyses of secondary outcomes (i.e. cost effectiveness, win odds analysis and long-term outcomes).

### 2.7 Timing of Outcome Assessments

Patients are followed for 2 years post-surgery, with continued follow-up via the LROI for long-term implant survival and mortality. All outcome assessment time points are described below. The outcomes are described in more detail in chapter 6 of this SAP.

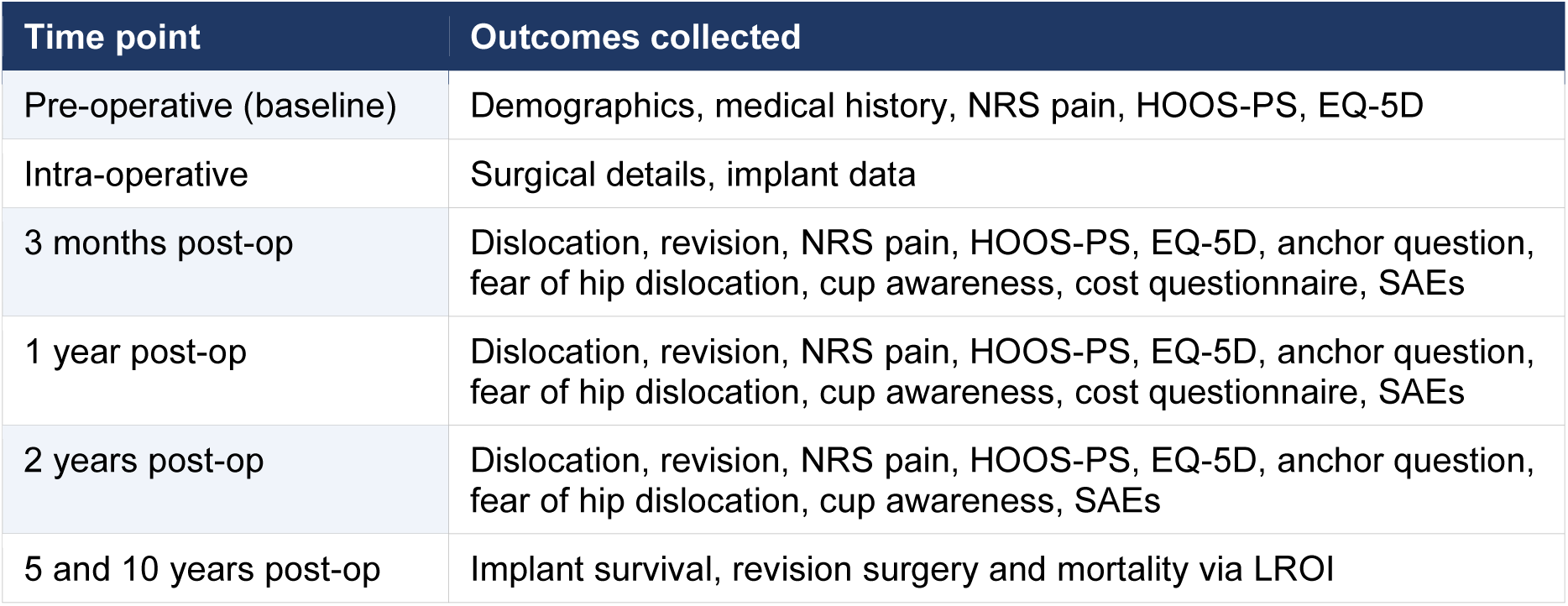

| Time point | Outcomes collected |
|---|---|
| Pre-operative (baseline) | Demographics, medical history, NRS pain, HOOS-PS, EQ-5D |
| Intra-operative | Surgical details, implant data |
| 3 months post-op | Dislocation, revision, NRS pain, HOOS-PS, EQ-5D, anchor question, fear of hip dislocation, cup awareness, cost questionnaire, SAEs |
| 1 year post-op | Dislocation, revision, NRS pain, HOOS-PS, EQ-5D, anchor question, fear of hip dislocation, cup awareness, cost questionnaire, SAEs |
| 2 years post-op | Dislocation, revision, NRS pain, HOOS-PS, EQ-5D, anchor question, fear of hip dislocation, cup awareness, SAEs |
| 5 and 10 years post-op | Implant survival, revision surgery and mortality via LROI |

## 3. Statistical Principles

### 3.1 Confidence Intervals and P-values

Results will be presented with 95% confidence intervals (95% CIs). P-values are reported only for the primary outcome, or upon request. A two-sided 5% significance level (α = 0.05) is considered, unless stated otherwise. Exact p-values will be reported to three decimal places; p-values below 0.001 will be reported as <0.001.

### 3.2 Multiplicity

The primary outcome (difference in number of patients who suffer a hip dislocation within 1 year after primary THA) is the single confirmatory hypothesis. Since 1 interim analysis is performed, the α-value in the primary analysis is adjusted to 0.0492 (see sections 3.5 and 6.1.2). No adjustment for multiplicity is made for secondary outcomes. The win odds analysis is a pre-specified additional analysis and is not subject to multiplicity correction. Results of exploratory analyses will be clearly labelled as such.

### 3.3 Adherence and Protocol Deviations

#### Adherence

Adherence is defined as receipt of the allocated cup type, using a posterolateral surgical approach by a surgeon participating in the trial. Participating hospitals and surgeons can use the implants of the industries they usually work with, as this study does not investigate a specific product, but rather the concept of dual mobility cups.

#### Protocol Deviations

Surgeons may deviate from the randomization scheme based on intra- operative findings; all deviations will be recorded as protocol deviations. Major deviations include wrong cup type placed (dual mobility instead of unipolar cup and vice versa), presence of exclusion criteria not caught at screening and not using the posterolateral surgical approach. Minor deviations include subtle variations of the posterolateral surgical approach and surgery performed by a surgeon not participating in the trial. All protocol deviations will be summarized by type and treatment arm.

### 3.4 Analysis Populations

#### Intention-To-Treat population (ITT)

In this population, patients are analyzed in the group they were randomized to, regardless of the type of cup they received and the surgical approach used. The primary analysis is performed in this ITT population as these randomized groups ensure minimal bias in estimating the treatment effect.

#### As-Treated population (AT)

In this population, patients are analyzed according to the type of cup they received, regardless of their randomized allocation. I.e. patients who were randomized to a DM cup but instead received a UP cup will then be assigned to the UP cup group, and vice versa.

Patients operated via any surgical approach are included. This analysis reflects the real-world effect of each cup type as actually implanted and will be reported as a sensitivity analysis. Note that this AT analysis is susceptible to bias, as crossovers may not be randomly distributed across treatment groups.

#### Per-Protocol population (PP)

In this population, only patients who received their allocated type of cup with a posterolateral surgical approach are included. Patients who received a different type of cup than randomly allocated to and/or underwent surgery with another surgical approach are excluded. If no data is available regarding the surgical approach, we assume that a posterolateral approach was used, as this was an inclusion criterion for the study. This PP approach will be reported as sensitivity analysis. The sample size -and therefore the statistical power – for this analysis will be lower than for the ITT analysis. Moreover, this PP analysis may introduce sources of bias, as protocol deviations may not be randomly distributed across treatment groups.

### 3.5 Blinding of treatment allocation

Patients were blinded for group allocation until after completion of the final questionnaire at 2-year follow-up (see Section 3.7). Participating surgeons and site investigators were not blind to treatment allocation. Data analysts will be blinded for group allocation.

To minimize analytical and interpretation bias, the outcome analyses will be conducted in separate blinded analytical phases. This sequential blinded analysis strategy is intended to preserve the objectivity of outcome analyses in the presence of a potentially large treatment effect on the primary endpoint (i.e. multiple dislocations in the UP group versus no or very few dislocations in the DM group). During each phase, treatment allocation will be coded as Group A and Group B, and the treatment identity will remain concealed from the trial statisticians and investigators involved in the analyses.

The primary analysis (chapter 6.1) of dislocation within one year will be performed using a dataset restricted to the variables required for the predefined primary analysis, excluding implant-specific identifiers or other variables that could reveal treatment allocation. The statistical code, analysis outputs, and written interpretation of the primary outcome will be finalized and locked before treatment allocation is unblinded for the primary analysis.

The secondary analyses (chapter 6.2) and safety analysis (chapter 6.3) will subsequently be analyzed in a separate blinded analytical phase using an independently prepared dataset. The statistical analysis plan, programming code, model specifications, and interpretation for these outcomes will likewise be finalized before treatment allocation is revealed.

### 3.6 Missing Data

Efforts are made to prevent missing data by sending reminders and making phone calls when appropriate. The level of missing data will be described in the final report, including reasons and patterns (by time point and treatment arm).

#### Primary outcome (dislocation)

This is a clinical event recorded in medical records, our questionnaires and the LROI. Missing data for the primary outcome is not anticipated. If no evidence of dislocation is found in any of the data sources (i.e. study questionnaires, medical records of the participating hospitals, and the LROI), we feel confident to assume that no dislocation occurred.

Therefore, no imputation will be applied to the primary outcome.

#### PROM scores

Our mixed-model analyses will account for missing data using maximum likelihood estimation. In the event of unforeseen numbers of missing values, a state-of-the-art solution will be sought in consultation with a statistician.

The extent pattern of missing data will be explored (MCAR/MAR/MNAR) using Little’s test and logistic regression analyses on missingness indicators.

#### Covariates

The adjusted primary analysis (see section 6.1.2) includes age, sex, American Society of Anesthesiologists (ASA), and body mass index (BMI). We expect little missing data on these covariates, as they are recorded both by the participating centers and by the LROI. If more than 5% of values remain missing, we will handle missing data with multiple imputation by chained equations, assuming data are missing at random.

## 4. Trial Population

### 4.1 Screening Data

All patients at the outpatient clinics of participating hospitals that meet the criteria to undergo elective primary THA will be screened for the in- and exclusion criteria (see section 5.2). To increase the generalizability of the results, the inclusion criteria were formulated as widely as possible. We will report the proportion of patients included in the study relative to the number of patients screened to characterize the representativeness of the trial sample relative to the whole population. The number of patients screened, included, and excluded will be presented in a flow diagram (Figure 1). Reasons for exclusion at screening and after randomization will be reported separately.

**Figure 1:**
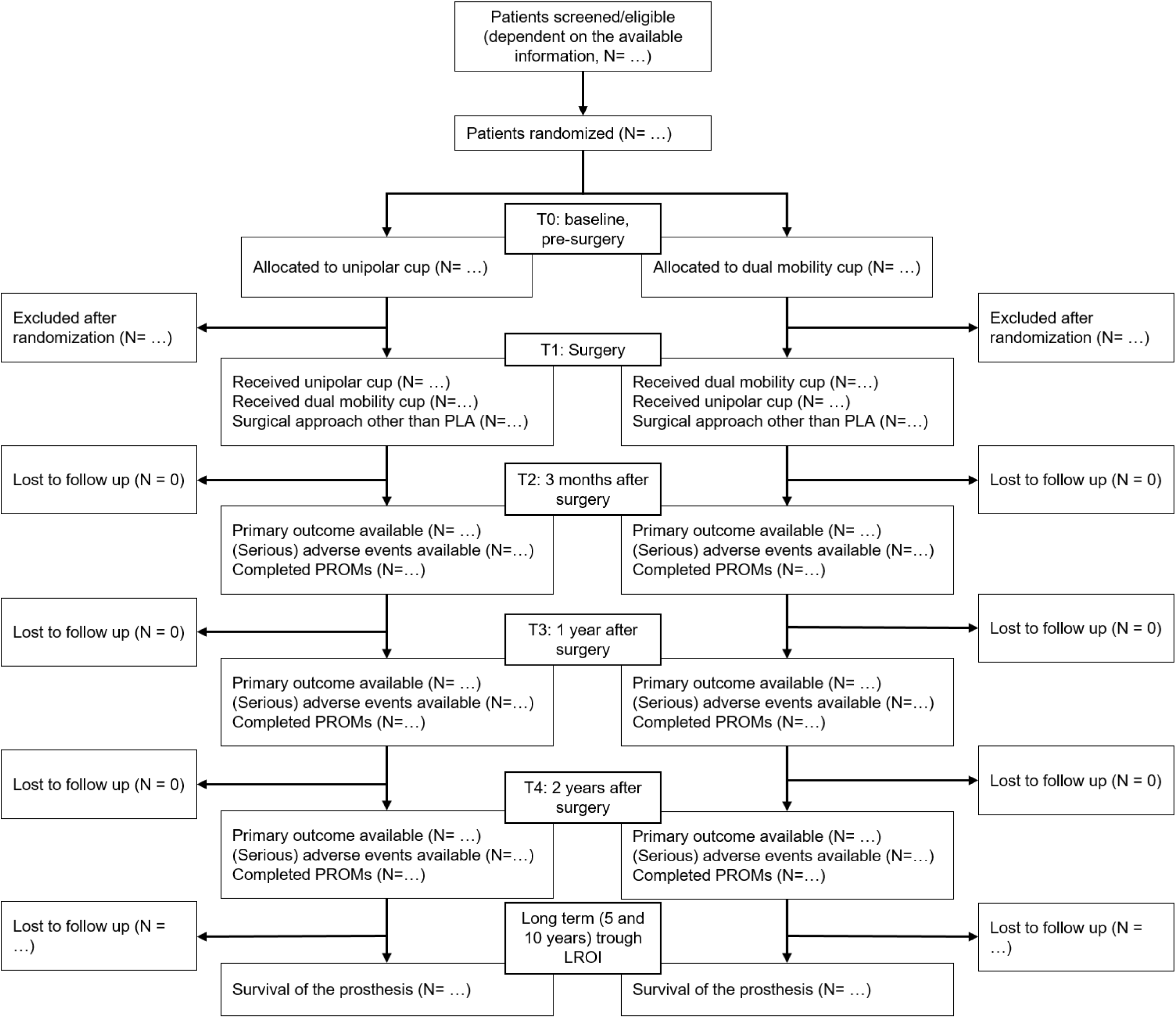
Flow diagram

### 4.2 Eligibility

To participate in the study, a patient must meet all the following criteria:

#### Inclusion criteria

- Patients eligible for elective primary THA with a 32mm or 36mm liner, for any indication
- THA using a posterolateral surgical approach
- Patients ≥ 70 years old
- Adequate comprehension of written and spoken Dutch or Spanish (for patients in the Vall d’Hebron University Hospital)

Patients meeting any of the following criteria are excluded from participation in the study:

#### Exclusion criteria

- Patients unable to complete PROMs
- Patients with dementia, epilepsy, spasticity, mental retardation, or alcoholism (If dementia or mental retardation is not already mentioned in the medical chart, this can be determined by doctors’ opinion)
- Patients not eligible for either a unipolar or dual mobility cup

### 4.3 Recruitment

A CONSORT flow diagram will be presented showing number of patients screened for eligibility, number of patients randomized, number of patients allocated to each group, number of patients who received allocated treatment, number of patients lost to follow-up with reasons, number of patients analyzed for the primary outcomes and number of patients in each analysis population.

### 4.4 Withdrawal and Follow-up

Patients may withdraw from the study at any time without consequence. Furthermore, the investigator can decide to withdraw a subject from the study for urgent medical or other reasons. Data collected prior to withdrawal will be included in analyses unless the patient also withdraws consent for data use. Withdrawal data will be summarized including timing of withdrawal, reasons (where provided), and group allocation.

### 4.5 Baseline Patient Characteristics

Participant demographics, surgical characteristics and baseline PROM scores will be summarized descriptively by treatment arm (See table 1, table 2 and table 3). Continuous variables will be summarized as mean (SD) or median (IQR) and compared using standardized mean differences (not p-values) to assess baseline balance. Categorical variables will be summarized as n (%). No formal significance testing will be performed on baseline characteristics.

**Table 1:** Participant demographics

| **Participant demographics** | **Dual mobility group (N = …)** | **Unipolar group (N = …)** |
|---|---|---|
| Age (years), mean (SD) |  |  |
| Sex, n (%) |  |  |
| Male |  |  |
| Female |  |  |
| BMI (kg/m<sup>2</sup>), mean (SD) |  |  |
| ASA classification, n (%) |  |  |
| ASA I |  |  |
| ASA II |  |  |
| ASA III |  |  |
| ASA IV |  |  |
| Smoking, n (%) |  |  |
| Yes |  |  |
| No |  |  |
| Unknown |  |  |
| Charnley classification, n (%) |  |  |
| A |  |  |
| B1 |  |  |
| B2 |  |  |
| C |  |  |
| Prior surgery to the affected joint, n (%) |  |  |
| Yes |  |  |
| No |  |  |

**Table 2:** Surgical characteristics

| Surgical characteristics | Dual mobility group (N = …) | Unipolar group (N = …) |
|---|---|---|
| Cup type, n (%)<br>Dual Mobility<br>Unipolar |  |  |
| Cup diameter (mm), mean (SD) |  |  |
| Femoral head diameter (mm), mean (SD) |  |  |
| Surgical time (minutes), mean (SD) |  |  |
| Surgical approach used, n (%)<br>Posterolateral<br>Direct superior<br>Anterolateral<br>Direct lateral<br>Posterior |  |  |
| Direct anterior |  |  |
| Surgical fixation, n (%)<br>Cemented<br>Cementless<br>Hybrid: acetabulum cemented |  |  |
| Cup brand, n (%)<br>Zimmer Biomet<br>Johnson % Johnson<br>LinkLima<br>Stryker<br>Medcontech<br>Surgival<br>Smith % Nephew<br>United Orthopedic Corporation<br>Other |  |  |
| Femoral Head brand, n (%)<br>Zimmer Biomet<br>Johnson % Johnson<br>LinkLima<br>Stryker<br>Medcontech<br>Surgival<br>Smith % Nephew<br>United Orthopedic Corporation<br>Other |  |  |

**Table 3:** Baseline PROM scores

| **Baseline PROM scores** | **Dual mobility group (N = …)** | **Unipolar group (N = …)** |
|---|---|---|
| NRS for pain in rest (0–10; higher = more pain), mean (SD) |  |  |
| NRS for pain during weight bearing (0–10; higher = more pain), mean (SD) |  |  |
| HOOS-PS total score (0–100; higher = better function), mean (SD) |  |  |
| EQ-5D utility index score (0–1; higher = better QoL), mean (SD) |  |  |
| EQ-5D VAS score (0-100; higher = better overall health), mean (SD) |  |  |

## 5. Analysis of Outcomes

This chapter describes all analyses planned for the trial’s primary publication. Analyses intended for future publications are described in Chapter 7. Unless stated otherwise, all analyses will be performed in the ITT population. The primary analysis will be repeated in the AT and PP populations as sensitivity analyses.

### 5.1 Primary outcome

#### 5.1.1 Outcome definition

| Outcome | Definition | Outcome ascertainment |
|---|---|---|
| Dislocation within 1 year | The number of patients who suffer a periprosthetic dislocation of the hip within 1 year after primary THA via a posterolateral approach with a dual mobility cup compared to a unipolar cup in patients ≥70 years old. | A dislocation event is ascertained when treated with either closed reduction (repositioning of the femoral head into the acetabular component by external manipulation of the limb, without a surgical incision) or open reduction (surgical repositioning of the femoral head into the acetabular component, via re-exposure of the joint). Ascertainment is based on medical records, study questionnaires, and the LROI. |

#### 5.1.2 Primary Outcome Analyses

The primary outcome (dislocation within 1 year) will be analyzed using the chi-square test. If the assumption of expected cell frequencies ≥5 is not met, Fisher’s exact test will be used instead. The significance level is set at α = 0.0492 to adjust for an increased type 1 error rate resulting from repeated hypothesis testing of accumulating data, introduced by our interim analysis. The absolute effect will be quantified as the proportion of patients who suffer a dislocation within 1 year in each group and the risk difference (the difference in the proportion of patients with a hip dislocation between groups) with a 95% CI. The relative effect will be quantified as the odds ratio (OR) with a 95% CI. If one group has zero events, the relative effect may not be estimable. In that case, the observed proportions and risk difference with corresponding 95% confidence interval will be reported as the final result. No continuity correction will be applied, as small expected counts can render corrected estimates unstable and difficult to interpret.

To adjust for potential confounding and clustering of patients within hospitals, we will use a multilevel logistic regression model (binomial) in R (version 4.5.2) as the pre-specified adjusted primary analysis. The model will include treatment allocation and the following covariates as fixed effects: age, sex, American Society of Anesthesiology classification (ASA) and Body Mass Index (BMI), and hospital as a random effect. These covariates were selected a priori based on established dislocation risk factors, as confirmed by our collaborators. The covariate adjusted treatment effect will be expressed as an OR with 95% CI and two-sided p-value. Given the anticipated small number of dislocations, the multilevel logistic regression model may fail to converge (i.e. may be unable to produce stable parameter estimates, or may produce estimates with excessively wide confidence intervals, reflecting insufficient events in the data to reliably estimate all model parameters). If the standard multilevel logistic regression model does not converge, we will refit the model with a reduced set of covariates, retaining treatment allocation, the random hospital intercept, and the covariates considered essential a priori: BMI and ASA classification. Age and sex will be removed from the model, in that order. If the reduced model also fails to converge, we will use Firth’s penalized logistic, incorporating a random hospital effect if feasible.

#### Update after discussing the blinded results, prior to unblinding of treatment allocation

The primary outcome analysis revealed zero events in one of the treatment groups, consistent with the anticipated low event rate. The implications for the statistical analysis were discussed with the supervising statistician. The interpretation of the results was subsequently discussed with the collaborator group (collaborators meeting on 20-10-2026).

Given the zero count in one of the groups, the OR cannot be estimated as a finite value. The primary outcome will therefore be reported as the observed proportion of dislocations in each group and the absolute difference in proportions between groups, with 95% confidence intervals calculated using methods appropriate for low event rates. Specifically, 95% confidence interval of individual proportions will be estimated using the Wilson score method and those of the absolute difference in proportions between groups using Newcombe’s method. If the between-group difference is statistically significant, we will also report the number needed to treat (NNT) to prevent one dislocation.

Due to zero events in one treatment group, the proposed multilevel logistic regression model was not considered appropriate because of complete separation, which hampers reliable estimation of the treatment effect. Firth’s penalized logistic regression will therefore be used for the final adjusted analysis. This model cannot account for clustering within hospitals. Given the limited total number of events, the model will be restricted to the a priori selected most relevant covariates: BMI and ASA classification.

### 5.2 Secondary outcomes

#### 5.2.1 Outcome definitions

| Outcome | Definition | Time point(s) |
|---|---|---|
| Total number of dislocations | The total number of dislocations within 1 year (including recurrent dislocations within patients) | Within 1 year after primary THA |
| HOOS-PS | Hip disability and Osteoarthritis Outcome Score Physical Short form total score (0–100; higher = better function). Validated Dutch version. | Baseline, 3 months, 1 year |
| EQ-5D index score | EuroQol 5 Dimensions utility index (0–1; higher = better QoL). Dutch reference values used for utility calculation. | Baseline, 3 months, 1 year |
| EQ-5D VAS | EQ-5D visual analogue scale for overall health (0–100). | Baseline, 3 months, 1 year |
| NRS pain | Numeric Rating Scale for pain in rest and during weight bearing (0–10; higher = more pain). | Baseline, 3 months, 1 year |

#### 5.2.2 Secondary Outcome Analyses

##### Total number of dislocations

The difference in the total number of hip dislocations within 1 year will be analyzed using generalized linear mixed model in R (version 4.5.2), consistent with the modelling framework for the adjusted primary analysis. The model will include treatment allocation as fixed effect and hospital as a random effect, with a negative binomial distribution and log link function. The negative binomial distribution was chosen because recurrent dislocations in a subset of patients may result in overdispersion of this count outcome variable. If the model does not converge, total event counts and crude event rates will be reported descriptively by group instead.

##### Patient-reported outcome measures (PROMs)

HOOS-PS, EQ-5D index score, EQ-5D VAS, and NRS pain will be analyzed using linear mixed- effects models with time, treatment × time interaction, and baseline PROM score as fixed effects, and patient and hospital as random effects (10). Results are expressed as adjusted mean difference with 95% CI. Distributional assumptions (normality of residuals) will be checked using Q-Q plots. If assumptions are not met, a rank-based or log-transformed analysis will be used as an alternative method.

*Update after discussing the blinded results, prior to unblinding of treatment allocation:*

Due to zero events in one treatment group, the generalized linear mixed model with negative binomial distribution failed to converge. As pre-specified, the total number of dislocations will therefore be reported descriptively as total event counts and crude event rates per group, without further statistical testing.

### 5.3 Safety analyses

#### 5.3.1 Outcome definitions

| **Outcome** | **Definition** | **Time point(s)** |
|---|---|---|
| Hip dislocation | Cumulative incidence of dislocation of the hip | Within 1 year after primary THA |
| Revision surgery | Cumulative incidence of revision surgery for any prosthetic component for any reason. | Within 1 year after primary THA |
| Periprosthetic joint infection | Cumulative incidence of infection of the prosthetic hip joint (regardless of the required treatment for the infection) | Within 1 year after primary THA |
| Mortality | Cumulative incidence of all-cause mortality. | Within 1 year after primary THA |

#### 5.3.2 Safety Analyses

All adverse events (AEs) and serious adverse events (SAEs) will be summarized as incidence rates, presented as n (%) by treatment arm. These adverse events are obtained from the clinical records, patient questionnaires and the LROI. The cumulative incidence of dislocation, revision surgery, prosthetic joint infection and mortality within 1 year will each be visualized separately using the Kaplan-Meier estimator. For dislocation, revisions surgery and prosthetic joint infection we will separately perform competing risk analysis, treating death as the sole competing risk. The time-to- event input for the Kaplan-Meier curve for these outcomes is based on the competing risk analysis. Dislocation, re-operation and prosthetic joint infection are not treated as competing risks for each other. Mortality will be visualized using the standard Kaplan-Meier estimator, as death has no competing risk in this context.

##### Update after discussing the blinded results, prior to unblinding of treatment allocation

During the collaborators meeting on 20-10-2026, the definition of the revision surgery outcome variable in the safety analyses was revised, prior to unblinding of treatment allocation. From a patient perspective, any reoperation on the hip represents a clinically meaningful event, regardless of whether prosthetic components are exchanged. The outcome will therefore be broadened to include all reoperations on the hip, comprising revision surgery (with component exchange), but also open reduction for dislocation, debridement, antibiotics and implant retention (DAIR) without component exchange, and any other surgical intervention in the hip joint.

### 5.4 Subgroup Analyses

The following pre-specified subgroup analyses will be conducted for the primary outcome (dislocation rate at 1 year):

- BMI (<30 vs. ≥30)
- Age group (70–79 years vs. ≥80 years)
- Sex (male vs. female)
- ASA classification (I–II vs. III–IV)

Higher age, female sex, higher BMI and higher ASA classification are established risk factors for dislocation after total hip arthroplasty (11). Patients at higher risk of dislocation may benefit more from the increased stability that a dual mobility cup provides. To explore this, we will assess effect modification by adding an interaction term between treatment allocation and each of these factors.

All subgroup analyses are exploratory. Subgroup effects will be assessed by including interaction terms (treatment × subgroup) in the primary analysis model — either the multilevel logistic regression model or Firth’s penalized logistic regression with a random hospital effect – depending on the used model. Each of the pre-specified subgroups will be tested individually, in a separate model, rather than including all interaction terms simultaneously. Results will be presented as forest plots with ORs and 95% CIs per subgroup. No adjustment for multiple comparisons will be made. Furthermore, randomization is not stratified by these subgroups (only by hospital), and the number of events is anticipated to be low, so we will interpret these results with caution.

#### Femoral head diameter

The use of a larger femoral head has been suggested to reduce the risk of post-operative dislocation (12). Femoral head diameter is inherently linked to cup type (UP or DM) and therefore cannot simply be included as an interaction term (treatment x subgroup) in the primary analysis model. Therefore, as a pre-specified subgroup analysis, the 1-year dislocation risk will be compared across three groups: unipolar cups >32 mm, unipolar cups ≤32 mm, and dual mobility cups. An overall comparison across the three groups will be performed using Chi square or Fisher-Freeman-Halton exact test (3×2 table). If warranted, pairwise comparisons between groups will be reported descriptively as proportions with 95% confidence intervals, without formal correction for multiple comparisons. Given the anticipated small number of events, results will be interpreted with caution and considered hypothesis-generating rather than confirmatory.

#### Update after discussing the blinded results, prior to unblinding of treatment allocation

Due to the limited number of total events and zero events in one treatment group, formal interaction testing is not feasible for any of the pre-specified subgroup analyses. All subgroup analyses will therefore be reported descriptively, presenting the number and proportion of patients with a dislocation in each subgroup with 95% confidence intervals calculated using the Wilson score method.

## 7 Objectives to be detailed in later versions of this SAP

### 7.1 Cost-effectiveness analysis

A cost-effectiveness analysis will be conducted from the societal perspective in accordance with Dutch guidelines (Zorginstituut Nederland). Costs will include direct healthcare costs (hospitalization, surgery, revision, treatment of dislocation) and indirect costs (patient time, productivity loss).

Uncertainty around incremental cost-effectiveness ratios (ICERs) will be expressed using bootstrapped cost-effect pairs plotted on cost-effectiveness planes. Cost-effectiveness acceptability curves will present the probability of dual mobility cups being cost-effective across a range of willingness-to-pay thresholds. Sensitivity analyses will test key assumptions.

### 7.2 Win Odds analysis

The win odds analysis evaluates a hierarchical composite of multiple outcomes, ranked by clinical importance. The included outcomes are all-cause mortality at 1 year, revision surgery within 1 year, dislocation within 1 year, prosthetic joint infection within 1 year, quality of life at 1 year (EQ-5D index score), physical functioning at 1 year (HOOS-PS total score) and pain at rest and pain during activity (Numeric Rating Scale for Pain) at 1 year. All-cause mortality is included as the most important in the order to account for the competing risk of death and prevent immortal time bias. The order of importance for the other outcomes is determined in a patient-centered way, with participants in the REDEP trial receiving an online questionnaire regarding the outcomes they consider most important. These outcomes are all-cause mortality, revision surgery, dislocation, prosthetic joint infection, quality of life (EQ-5D index score), physical functioning (HOOS-PS total score), pain at rest and pain during activity (Numeric Rating Scale for Pain). The hierarchy of the outcomes is defined by patients.

### 7.3 Outcomes at two year follow up

The results at 2 years after surgery (dislocations, PROMs and safety outcomes) will be analyzed and reported similarly to the results at 1 year after surgery, as detailed in this SAP.

### 7.4 Long-term (5-10 year) implant survival

Time to first revision surgery will be analyzed using Kaplan-Meier survival estimation and a log-rank test. The hazard ratio will be estimated using a multilevel Cox proportional hazards model adjusted for the same covariates as the primary analysis. Proportional hazards assumption will be verified using Schoenfeld residuals.

### 7.5 Other outcomes for future analyses

These outcomes will not be included in the trial’s primary publication but will be addressed in future analyses and subsequent versions of this SAP.

| Outcome | Definition | Time point(s) |
|---|---|---|
| Anchor question | 7-point Likert scale regarding overall change in physical functioning (1 = Deteriorated significantly, 7 = Improved significantly) | 3 months, 1 year, 2 years |
| Fear of hip dislocation | 5-point Likert scale (1 = not afraid, 5 = very afraid). | 3 months, 1 year, 2 years |
| Awareness of cup type | Whether patient is aware of assigned cup type (yes/no). If yes, source of information. | 3 months, 1 year, 2 years |
| Cup size | The size of the cup may affect the risk of dislocation, which was not specified in the original protocol, but may be interesting for a post hoc analysis. | During surgery |
| Leg length | Leg length difference after total hip replacement | Post-operative X-ray and/or clinical consultation |

## **8** Statistical Software

All analyses will be performed in SPSS (version 29.0) and R (version 4.5.2).

## Data Availability

All statistical analyses described in the statistical analysis plan will be reported in a manuscript and will be published in a scientific journal with open access.

## List of Abbreviations

| **Abbreviation** | **Definition** |
|---|---|
| AE | Adverse Event |
| ASA | American Society of Anaesthesiology classification |
| AT | As Treated |
| BMI | Body Mass Index |
| CI | Confidence Interval |
| CONSORT | Consolidated Standards of Reporting Trials |
| EQ-5D | EuroQol 5 Dimensions |
| HOOS-PS | Hip disability and Osteoarthritis Outcome Score Physical Short form |
| ICH | International Conference on Harmonisation |
| IQR | Interquartile range |
| ITT | Intention To Treat |
| LROI | Landelijke Registratie Orthopedische Interventies (Dutch Arthroplasty Register) |
| NRS | Numeric Rating Scale |
| OR | Odds Ratio |
| PP | Per Protocol |
| PROM | Patient Reported Outcome Measure |
| QALY | Quality Adjusted Life Year |
| SAE | Serious Adverse Event |
| SAP | Statistical Analysis Plan |
| SD | Standard Deviation |
| THA | Total Hip Arthroplasty |
| WMO | Wet Medisch-wetenschappelijk Onderzoek met Mensen |

